# Saliva swabs are the preferred sample for Omicron detection

**DOI:** 10.1101/2021.12.22.21268246

**Authors:** Gert Marais, Nei-yuan Hsiao, Arash Iranzadeh, Deelan Doolabh, Annabel Enoch, Chun-yat Chu, Carolyn Williamson, Adrian Brink, Diana Hardie

**Affiliations:** Department of Medical Microbiology, University of Cape Town, Cape Town, South Africa; Department of Medical Virology, University of Cape Town, Cape Town, South Africa; Green Point Diagnostic Virology Laboratory, National Health Laboratory Service, Cape Town, South Africa

## Abstract

The Omicron variant is characterised by more than 50 distinct mutations, the majority of which are located in the spike protein. The implications of these mutations for disease transmission, tissue tropism and diagnostic testing are still to be determined. We evaluated the relative performance of saliva and mid-turbinate swabs as RT-PCR samples for the Delta and Omicron variants. The positive percent agreement (PPA) of saliva swabs and mid-turbinate swabs to a composite standard was 71% (95% CI: 53-84%) and 100% (95% CI: 89-100%), respectively, for the Delta variant. However, for the Omicron variant saliva and mid-turbinate swabs had a 100% (95% CI: 90-100%) and 86% (95% CI: 71-94%) PPA, respectively. This finding supports ex-vivo data of altered tissue tropism from other labs for the Omicron variant. Reassessment of the diagnostic testing standard-of-care may be required as the Omicron variant become the dominant variant worldwide.

## Introduction

SARS-CoV-2 variants are characterised by distinct mutations which impact on disease transmissibility, immune escape, diagnostics and possibly tissue tropism. Omicron, in particular, has an extraordinary number of mutations, with at least 50 mutations across the genome, 30 of which are located in the spike protein and 15 in the receptor binding domain.^1^ While functional change in terms of receptor binding is currently to be elucidated, the pattern of viral shedding and resulting impact on diagnostic sampling methods is currently unknown.

## Methods

As part of an on-going study^2^ to evaluate the diagnostic performance of different sample types, we recruited 382 acutely symptomatic, non-hospitalised patients who presented for SARS-CoV-2 testing between August and December 2021 at the Groote Schuur Hospital COVID testing centre in Cape Town. Paired mid-turbinate (MT) and saliva (SA) swabs were collected and tested by RT-PCR (Supplementary methods).

Samples were classified as Omicron or Delta based on whole genome sequencing data, diagnostic PCR target failures and sampling date (Supplementary methods).^1,3,4^ A composite standard for SARS-CoV-2 infection was used for comparison of sample types, with infection considered present if SARS-CoV-2 RNA was detected on either the MT or matched SA swab.

## Results

The positive percent agreement (PPA) of SA swabs and MT swabs to this standard was 71% (95% CI: 53-84%) and 100% (95% CI: 89-100%), respectively, for the Delta variant. This was similar to our previous findings for the Beta variant.^2^ However, for the Omicron variant SA and MT swabs had a 100% (95% CI: 90-100%) and 86% (95% CI: 71-94%) PPA, respectively (Supplementary Figure 1). The mean RT-PCR cycle threshold differences between MT and SA, using the nucleocapsid gene target as a reference, were 5.2 (SD ± 5.1, P<0.0001) and 1.5 (SD ± 5.9, P=0.18) for Delta and Omicron respectively. The median time from symptom onset to positive test for Delta and Omicron assigned cases was 3 days (range: 1-10) and 2 days (range: 0-7).

## Conclusion

These findings suggest that the pattern of viral shedding during the course of infection is altered for Omicron with higher viral shedding in saliva relative to nasal samples resulting in improved diagnostic performance of saliva swabs. This supports the ex-vivo finding of improved viral replication in upper respiratory tract tissue and possibly altered tissue tropism.^5^ This is an important finding as the current standard of care for diagnosis using swabs of the nasal or nasopharyngeal mucosa may be suboptimal for the Omicron variant.

## Supporting information

Supplementary raw data

## Data Availability

All data produced in the present study are available upon reasonable request to the authors.

## Acknowledgements

This research was funded in whole, or in part, by Wellcome [203135/Z16/Z]. For the purpose of Open Access, the author has applied a CC BY public copyright licence to any Author Accepted Manuscript version arising from this submission.

Testing was conducted at the Groote Schuur Hospital and Green Point National Health Laboratory Service diagnostic virology laboratories.

## Ethics statement

This research has been approved by the University of Cape Town Human Research Ethics Committee (Ref: 420/2020).

## Figures

**Figure 1.**
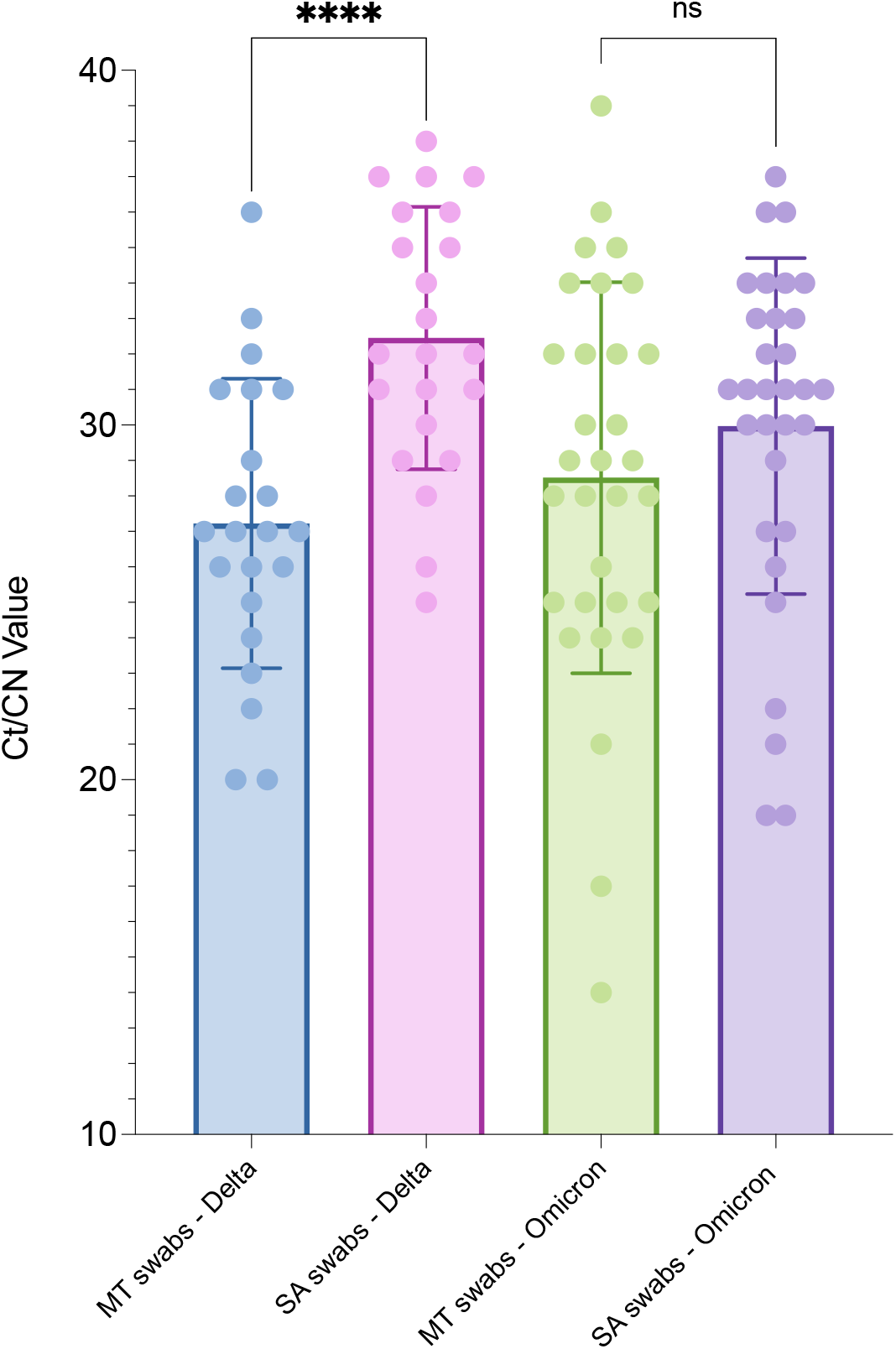
The cycle threshold (Ct) or cycle number (CN) values for paired mid-turbinate (MT) and saliva (SA) swabs are shown for Delta and Omicron variant positive samples. Paired samples were tested on the same diagnostic platform on the same day and samples where only the MT or SA swab was positive were excluded from the analysis. The nucleocapsid (N) gene Ct value was used for analysis if the sample was tested with the Allplex™ 2019-nCoV assay (Seegene, South Korea). This was because the Delta and Omicron variants are not associated with N gene target failure and other assays used also target the N gene. Statistical analysis consisted of paired t-tests performed using GraphPad Prism version 9.3.0 for macOS, GraphPad Software, San Diego, California USA, www.graphpad.com. The bar represents the mean Ct value with error bars showing 1 standard deviation. ns: not significant. ****: P value < 0.0001.

## Supplementary methods

### Swab collection

Swabs were self-collected by the study participants under supervision of a healthcare worker.

Participants should not have had any food, drink, tobacco or gum in the 30 minutes preceding saliva swab collection. Participants were initially instructed to cough 3-5 times, covering their mouths with the inner elbow. They were then asked to swab on the inside of both cheeks, above and below the tongue, on the gums and hard palate. A minimum swabbing duration of 30 seconds was required. The swab was transported in a sealed container to the laboratory without any transport media.

Mid-turbinate swabs were collected by a healthcare worker. The swab was inserted 2-3 cm into each nostril and transported in a sealed container to the laboratory without any transport media.

On arrival in the laboratory, all swabs were placed in 2 ml Sarstedt containers with 1.5 ml of sterile autoclaved 0.9% saline in preparation for downstream RT-PCR testing.

### RT-PCR

Swabs were tested by the Groote Schuur Hospital National Health Laboratory Service (NHLS) diagnostic virology laboratory in Cape Town, South Africa. The assays in used by this laboratory during the study period were the Allplex™ 2019-nCoV assay (Seegene, South Korea) (n=343), the Abbott RealTime SARS-CoV-2 assay (Abbott Laboratories, USA) (n=7) and the Abbott Alinity m SARS-CoV-2 assay (Abbott Laboratories, USA) (n=32). The assay used was based on laboratory operational requirements and no study-specific considerations or requirements were in place. The Abbott assay were run as per kit package inserts and subject to the operational requirements of a South African National Accreditation System (SANAS) accredited diagnostic virology laboratory. The Seegene assay was run with an in-house developed laboratory-specific sample processing technique which was subject to a validation as per SANAS requirements. Paired samples were in all cases tested using the same RT-PCR platform.

Selected samples (n=31) that tested positive primarily were assessed for Spike gene target failure using the TaqPath COVID-19 CE-IVD RT-PCR Kit (Thermo Fisher Scientific, USA) at the Green Point NHLS diagnostic virology laboratory.

### Variant classification

A confirmed classification as Delta or Omicron was based on whole genome sequencing as previously described.^1^ A probable assignment was based on variant-specific RT-PCR gene target failure profiles noted during diagnostic testing^3,4^ and a possible assignment was based on the local dominant circulating variant at the time of sample collection.^1^ RNA-dependent RNA-polymerase (RdRp) gene target failure (R-GTF) was considered present if the RdRp Ct value was >3.5 cycles greater than the Envelope (E) gene Ct value. In cases where the RdRp gene was not detected, R-GTF was considered present if the E gene had a Ct value of <30. Spike (S) gene target failure was considered present if all assay SARS-CoV-2 gene targets other than S were detected. The Delta variant was dominant in Cape Town prior to the 19^th^ of November 2021 and the Omicron variant subsequently (Supplementary Figure 1).^1^ For the purposes of positive percent agreement, negative percent agreement, positive predictive value and negative predictive value calculation a Delta or Omicron possible, probable or confirmed classification was accepted.

## Supplementary figures

**Supplementary Figure 1.**
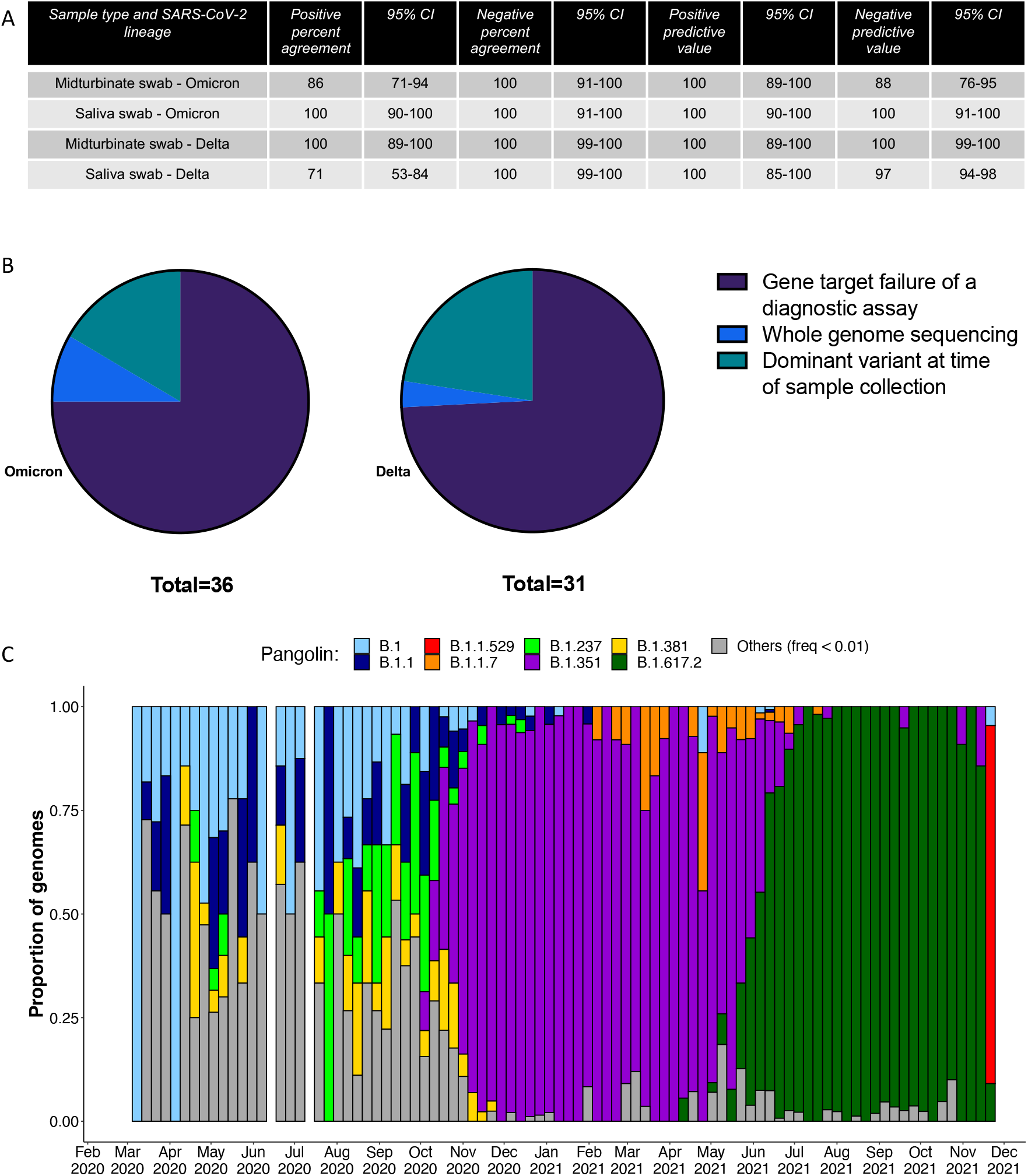
(A) Table showing the positive and negative percent agreement and positive and negative predictive values for mid-turbinate and saliva swabs with 95% confidence intervals shown. Confidence intervals were calculated using the Wilson-Brown method using GraphPad Prism version 9.3.0 for macOS, GraphPad Software, San Diego, California USA, www.graphpad.com. For the Delta variant, 277 samples tested negative, for 22 samples both the saliva (SA) and mid-turbinate (MT) swab tested positive and for 9 samples only the MT swab tested positive. No samples tested SA swab positive only. For the Omicron variant, 38 samples tested negative, for 31 samples both the SA and MT swab tested positive and for 5 samples only the SA swab tested positive. No samples tested MT swab positive only. (B) The proportions of SARS-CoV-2 lineage assignments by listed criteria for samples testing positive are shown. 36 samples were classified as Omicron, 75% as probable due to S gene target failure during diagnostic testing, 17% as possible due to the dominant circulating variant at the time of sample collection and 8% as confirmed by whole genome sequencing. Similarly, 31 samples were classified as Delta, 74% as probable due to RdRp gene target failure during diagnostic testing, 23% as possible due to the dominant circulating variant at the time of sample collection and 3% as confirmed by whole genome sequencing. (C) The longitudinal proportion of Pangolin lineages for samples originating in the Western Cape, South Africa.

